# Characteristics of 24,516 Patients Diagnosed with COVID-19 Illness in a National Clinical Research Network: Results from PCORnet^®^

**DOI:** 10.1101/2020.08.01.20163733

**Authors:** Jason P. Block, Keith A. Marsolo, Kshema Nagavedu, L. Charles Bailey, Henry Cruz, Christopher B. Forrest, Kevin Haynes, Adrian F. Hernandez, Rainu Kaushal, Abel Kho, Kathleen M. McTigue, Vinit P. Nair, Richard Platt, Jon Puro, Russell L. Rothman, Elizabeth Shenkman, Lemuel Russell Waitman, Mark G. Weiner, Neely Williams, Thomas W. Carton, on behalf of PCORnet, Network Partners

## Abstract

**Background:** National data from diverse institutions across the United States are critical for guiding policymakers as well as clinical and public health leaders. This study characterized a large national cohort of patients diagnosed with COVID-19 in the U.S., compared to patients diagnosed with viral pneumonia and influenza.

**Methods and Findings:** We captured cross-sectional information from 36 large healthcare systems in 29 U.S. states, participating in PCORnet^®^, the National Patient-Centered Clinical Research Network. Patients included were those diagnosed with COVID-19, viral pneumonia and influenza in any care setting, starting from January 1, 2020. Using distributed queries executed at each participating institution, we acquired information for patients on care setting (any, ambulatory, inpatient or emergency department, mechanical ventilator), age, sex, race, state, comorbidities (assessed with diagnostic codes), and medications used for treatment of COVID-19 (hydroxychloroquine with or without azithromycin; corticosteroids, anti-interleukin-6 agents).

During this time period, 24,516 patients were diagnosed with COVID-19, with 42% in an emergency department or inpatient hospital setting; 79,639 were diagnosed with viral pneumonia (53% inpatient/ED) and 163,984 with influenza (41% inpatient/ED). Among COVID-19 patients, 68% were 20 to <65 years of age, with more of the hospitalized/ED patients in older age ranges (23% 65+ years vs. 12% for COVID-19 patients in the ambulatory setting). Patients with viral pneumonia were of a similar age, and patients with influenza were much younger. Comorbidities were common, especially for patients with COVID-19 and viral pneumonia, with hypertension (32% for COVID-19 and 46% for viral pneumonia), arrhythmias (20% and 35%), and pulmonary disease (19% and 40%) the most common. Hydroxychloroquine was used in treatment for 33% and tocilizumab for 11% of COVID-19 patients on mechanical ventilators (25% received azithromycin as well).

**Conclusion and Relevance:** PCORnet leverages existing data to capture information on one of the largest U.S. cohorts to date of patients diagnosed with COVID-19 compared to patients diagnosed with viral pneumonia and influenza.

---

The novel coronavirus SARS-CoV-2 has resulted in unprecedented challenges to the global medical and public health sectors. The virus has spread rapidly across the world, leading policymakers, clinicians, first responders, and public health professionals to coordinate responses with limited information about effects on diverse populations and potential treatments. Over the first few months of the pandemic, data on patients with COVID-19 came from varied sources, often as single-institution or regional case series or through case reports to public health authorities, typically based on positive lab tests (1-5). Case reports to public health authorities and popular dashboards that rely on this information often lack detailed information about patient characteristics, use of treatments, or course of illness (2, 6-9).

A rapidly evolving pandemic presents tremendous uncertainty. The pattern of disease is typically unknown initially, and knowledge can flow in asymmetric ways, often driven by anecdotes, press reports, and social media. Throughout much of the COVID-19 pandemic, testing has been insufficient, limiting knowledge about the scope of the pandemic and characteristics of patients infected. Treatments have become mainstays before being fully vetted (10-12).

Leaders, institutions, researchers, and patients need extensive and accurate information about characteristics of patients infected, utilization of treatments and disease course, ideally from diverse settings across large geographic regions. Research networks that can capture this information across many healthcare systems have an important role during a pandemic response, especially because they can provide more generalizable, real-world information on a diverse population. In this article, we describe characteristics of one of the largest national cohorts of patients to date diagnosed with COVID-19 and compare this cohort to patients diagnosed with other viral illnesses in institutions participating in PCORnet^®^, the National Patient-Centered Clinical Research Network.

## Methods

Developed with funding by the Patient-Centered Outcomes Research Institute^®^ (PCORI^®^), PCORnet^®^ is a network-of-networks developed to facilitate comparative effectiveness research using electronic health record (EHR) and other healthcare data (13, 14). The Network currently has participation from 251 medical institutions, 337 hospitals, 3,574 primary care practices, and 1,024 community clinics embedded within nine Clinical Research Networks (i.e., consortia of health systems called PCORnet Network Partners) and 61 data contributing sites (i.e., individual health systems); the Network also has participation from two Health Plan Research Networks. PCORnet Network Partners (NPs) are well suited to develop and analyze cohorts of patients across many of the largest health systems in the United States. While set up to accommodate comparative effectiveness and epidemiologic research, the network has features that allow for a transition to rapid querying of data in a medical and public health crisis. In this study on COVID-19 patients, 37 of the 61 data contributing sites participated.

### PCORnet Common Data Model

PCORnet operates as a distributed research network, where data are held locally behind institutional firewalls, with queries sent to institutions from a central PCORnet Coordinating Center and results returned in the form of aggregate counts or summary statistics. NPs also have the capacity to generate patient-level data and to link electronic health records to claims data for specific projects. The NPs utilize the PCORnet Common Data Model, in which each data contributing site maps its clinical and/or administrative claims data into a standardized representation (15). The most recent version of the PCORnet Common Data Model, v5.1, provides standards for information on demographics, diagnoses, vital signs, laboratories, prescriptions and dispensed medications, medications administered (multiple settings), procedures, and facility and patient geographic information, among other information (15). These data are normally updated quarterly by sites, and the PCORnet Coordinating Center curates the data to ensure conformance with the Common Data Model, data plausibility, and data completeness (16). The Coordinating Center provides metrics based on these data quality domains to sites, allowing them to troubleshoot any data problems; some deficiencies require mitigation before the site can participate in PCORnet queries.

Prompted by the COVID-19 pandemic, PCORnet NPs agreed to abruptly change the operation of the network, allowing for rapid capture of information on patients infected with COVID-19. To facilitate this, starting in early April, NPs began updating their data weekly instead of quarterly. Rather than doing this for all patients (which can involve millions of patients and require considerable time), PCORnet sites focused the refresh on patients that had at least one International Classification of Diseases, 10^th^ revision, Clinical Modification (ICD-10-CM) diagnostic code for a respiratory illness or had a lab order for a SARS-CoV-2 test recorded beginning January 1, 2020 (**Fig 1**) (17). This respiratory illness filter created a COVID-19-focused Common Data Model that included all information available in the typical “full population” Common Data Model but for many fewer patients. The Common Data Model respiratory illness filter included ICD-10-CM codes for coronavirus (COVID-19 and other), pneumonia, bronchitis, influenza, upper respiratory infections, fever, cough, respiratory failure or acute respiratory distress syndrome, and shortness of breath (**S Methods Note**). The codes for SARS-CoV-2 laboratory orders were Special Use terms identified via the Logical Observation Identifiers Names and Codes (LOINC) Prerelease Page, as well as guidance from the Centers for Medicare and Medicaid Services (CMS) on SARS-CoV-2-related Healthcare Common Procedure Coding System (HCPCS) codes (18). Sites first began populating their COVID-19 Common Data Model during the week of March 27^th^. The timeline for implementation varied by site based on ability to change data infrastructure and alter governance to allow for frequent updates and queries.

**Fig 1.**
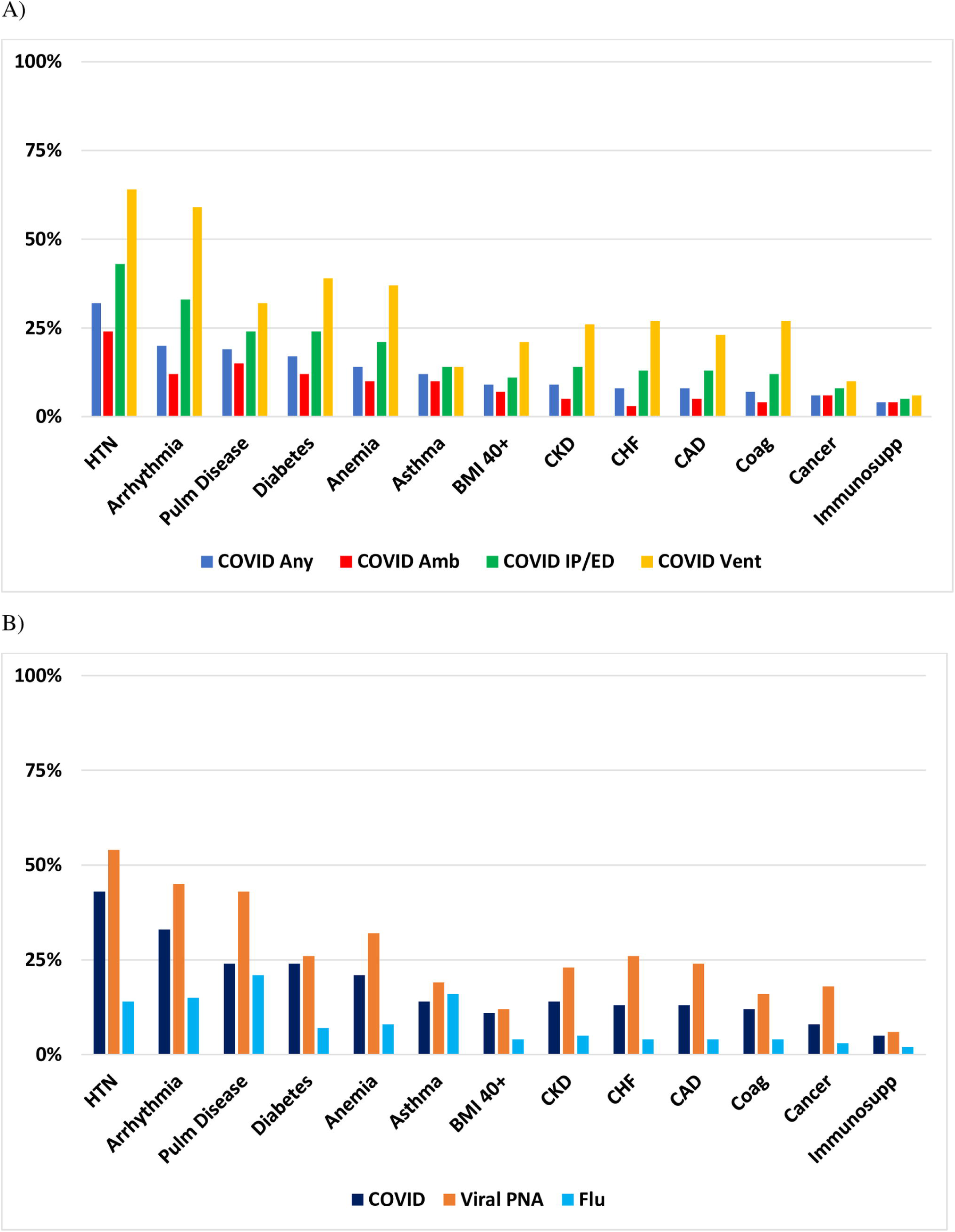
PCORnet Common Data Models, including standard and COVID-19 Common Data Model. The PCORnet Common Data Model includes information collected through various clinical data systems, including electronic health records (EHR) and health insurance claims or pharmacy dispensing records. Once data are entered by the user, systems transfer data to a data warehouse that is unique to their own health system and system vendor (e.g., Epic, Cerner). The typical process for populating the PCORnet Common Data Model is through an extraction-transfer-loading (ETL) procedure that allows for transformation of raw information into a form that is compliant with the Common Data Model specifications. This transformation allows for consistency across health systems and centralized querying of data, and the PCORnet ETL and Common Data Model data refresh occurs quarterly. The PCORnet COVID-19 Common Data Model utilizes a similar process but rather than including information on all patients, it includes information only on patients who have had at least one respiratory illness diagnostic code or a lab test ordered for SARS-CoV-2 since January 1, 2020. This process occurs weekly and includes data with a lag time between 1 day to 7 days, depending on the site.

### Query Development Process and Cohort Characterization

The PCORnet Coordinating Center has developed standardized statistical programs, called modular programs, that can capture aggregate descriptive information on defined cohorts. These programs define a cohort or multiple cohorts and then capture broad descriptive information, including demographics, comorbidities (and symptoms based on billing codes), prescriptions or hospital-administered medications, presence of laboratory results, and geographic information. NPs return data as counts and frequencies for variables specified; the PCORnet Coordinating Center combines results across NPs, producing a summary report of Network-wide aggregate results. Submission and use of these aggregate data are covered by a Network-wide Data Security Agreement.

Starting from the broader group of patients whose information was in the COVID-19 Common Data Model, we defined a cohort of patients with ICD-10-CM diagnostic codes for COVID-19 or other coronaviruses (our case definition for COVID-19 for this study), viral pneumonia, or influenza (**S1 Table)**. The case definition did not use laboratory data. We assessed patients with these three infections to enable some characterization of patients with other infections relevant to COVID-19 as comparison cohorts. The viral pneumonia and influenza cohorts excluded patients who had COVID-19 (or other coronavirus) ICD-10-CM codes. We stratified our cohort based on these three infections as well as the setting of diagnosis: 1) any setting, 2) ambulatory, 3) inpatient or emergency department, or 4) inpatient or emergency department with procedure codes identifying use of mechanical ventilation. Patients could be included in multiple care settings, if patients received diagnostic codes in more than one. We identified mechanical ventilation usage through ICD-10-CM, HCPCS and Current Procedural Terminology (CPT) codes. All codes for case definitions, comorbidities, treatments and labs were posted on GitHub (https://github.com/PCORnet-DRN-OC/Query-Details/tree/master/COVID_weekly_query/2020_04_29).

### Comorbidities, Treatments and Symptoms

We captured aggregate data across NPs for demographic characteristics of patients, including age group, sex, and race, using value sets defined in the PCORnet Common Data Model v5.1. Sex categories allowed for Male, Female, and Other. For race, several categories, including Native Hawaiian/Other Pacific Islander and Alaskan Native/American Indian, were less common and typically <1% for all cohorts.

We included a detailed assessment of comorbidities. We chose these based on discussions with experts regarding the comorbidities that might affect outcomes with COVID-19, as well as information presented in early publications regarding COVID-19 outcomes. Patients were considered to have a disease/condition if they had at least one ICD-10-CM code for the disease/condition recorded any time over a three-year period prior to the date of their index diagnosis. We required only one code to account for the possibility that some patients had their first encounter with the healthcare system for their admission for COVID-19 and may only have one code listed for a comorbidity. Using RxNorm and National Drug Codes (NDC) lists, we identified use of immunosuppressive medications over the same three-year period (19, 20). Similarly, we captured information on medications being used for treatment, with an assessment of prescriptions given in the 14-day period before and after the index diagnosis. These included hydroxychloroquine and chloroquine, alone or in combination with azithromycin, as well as corticosteroids and anti-IL-6 agents (siltuximab, sarilumab, tocilizumab) used to treat the cytokine storm that can occur with severe COVID-19. We also captured use of critical care procedure/billing CPTs (99291, 99292) during this period and the presence of laboratory tests, identified with LOINC codes, that are commonly used in the assessment of COVID-19. Lastly, we categorized weight status for each cohort, using the most recent height and weight available, up to one year prior to the index diagnosis date. We calculated measures separately by age group, using body mass index (BMI) for adults and Centers for Disease Control and Prevention macros for calculation of BMI z-scores and weight status for children < 20 years of age (21).

## Results

The query captured aggregate data on 24,516 patients with a COVID-19 diagnosis from 36 data contributing sites, across 29 states; 37 sites responded to the query, but 1 site had no cases reported (**S2 Table**). Across the 36 sites, percent contributions to the overall COVID-19 population ranged from <1% to 14% with 11 sites each contributing ≥ 4%; these 11 sites included 70% of all COVID-19 patients. The most commonly represented states were Illinois (18%), Wisconsin (11%), and Florida (9%), with 18% of patients missing geographic information. Among patients diagnosed with COVID-19, 42% were diagnosed in an emergency department or inpatient hospital setting, and the most commonly used codes for the diagnosis of COVID-19 were B34.2 (19%), B97.29 (33%), and U07.1 (62%). A lab test for SARS-CoV-2 was present for 68% of patients diagnosed (**S1 Figure**). For comparison conditions, the query identified 79,639 diagnosed with viral pneumonia (53% in hospital/ED) and 163,984 with influenza (41% in hospital/ED). Labs commonly used in the assessment of patients with COVID-19 were variably available in the PCORnet Common Data Model, with creatinine, alanine aminotransferase and aspartate aminotransferase, and lymphocyte count the most common.

Most COVID-19 patients were diagnosed in March and April 2020 (25% and 53% respectively), in contrast to viral pneumonia and influenza patients, who were most often diagnosed in January and February (64% and 84% respectively). Only 0.4% of influenza cases were diagnosed in April/May 2020 (**Table 1**). Among COVID-19 patients, 68% were 20 to <65 years of age, with more of the hospitalized/ED patients in older age ranges (23% 65+ years compared to 12% for COVID-19 patients in the ambulatory setting) (**Table 1**). Patients with influenza were younger than patients with COVID-19 or viral pneumonia (63% hospitalized/ED with influenza were <20 years of age compared to 16% for COVID-19 and 21% for viral pneumonia). More female patients (55%) received a COVID-19 diagnosis overall, with a predominance of men among those on a mechanical ventilator (57%).

**Table 1:**
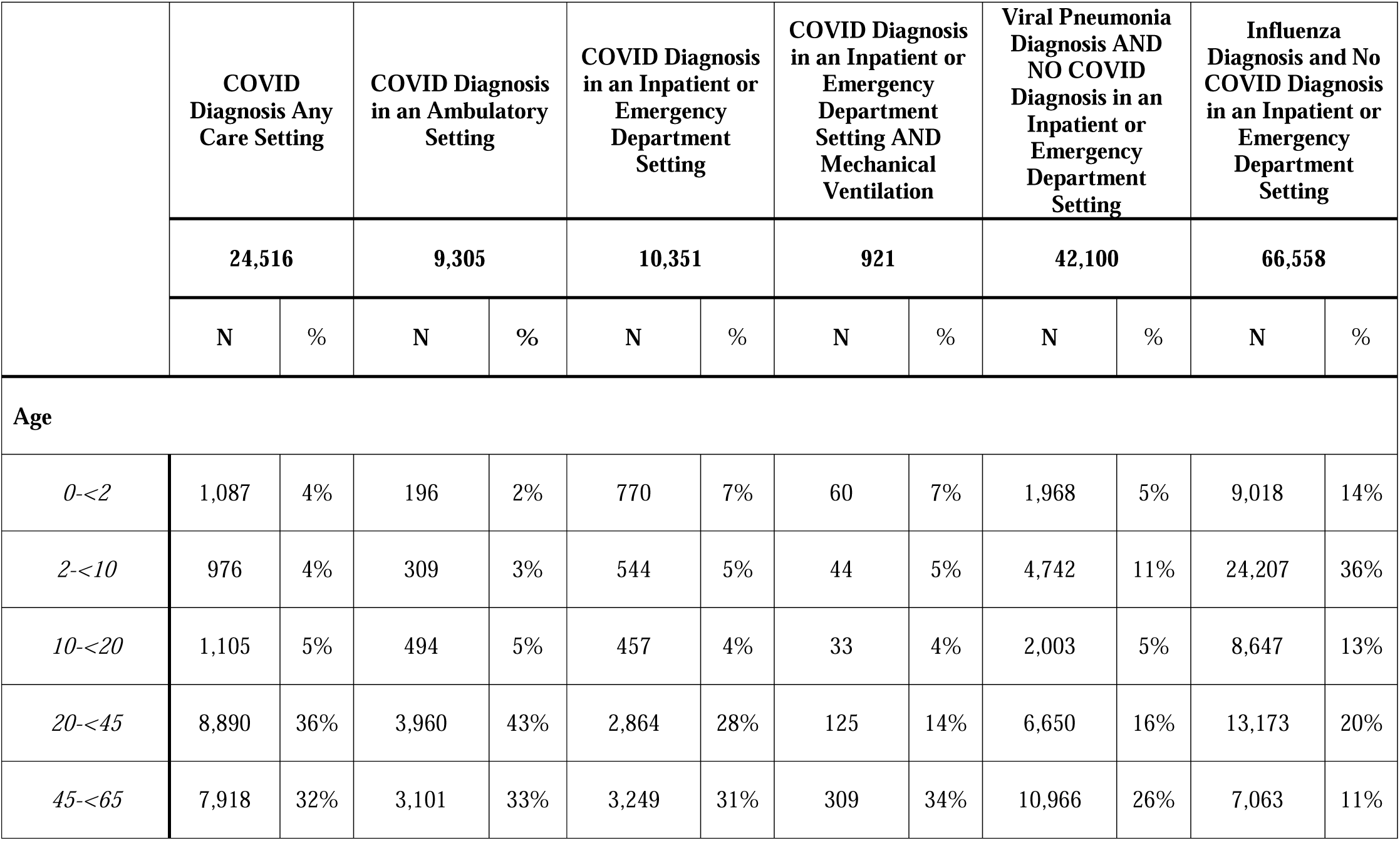
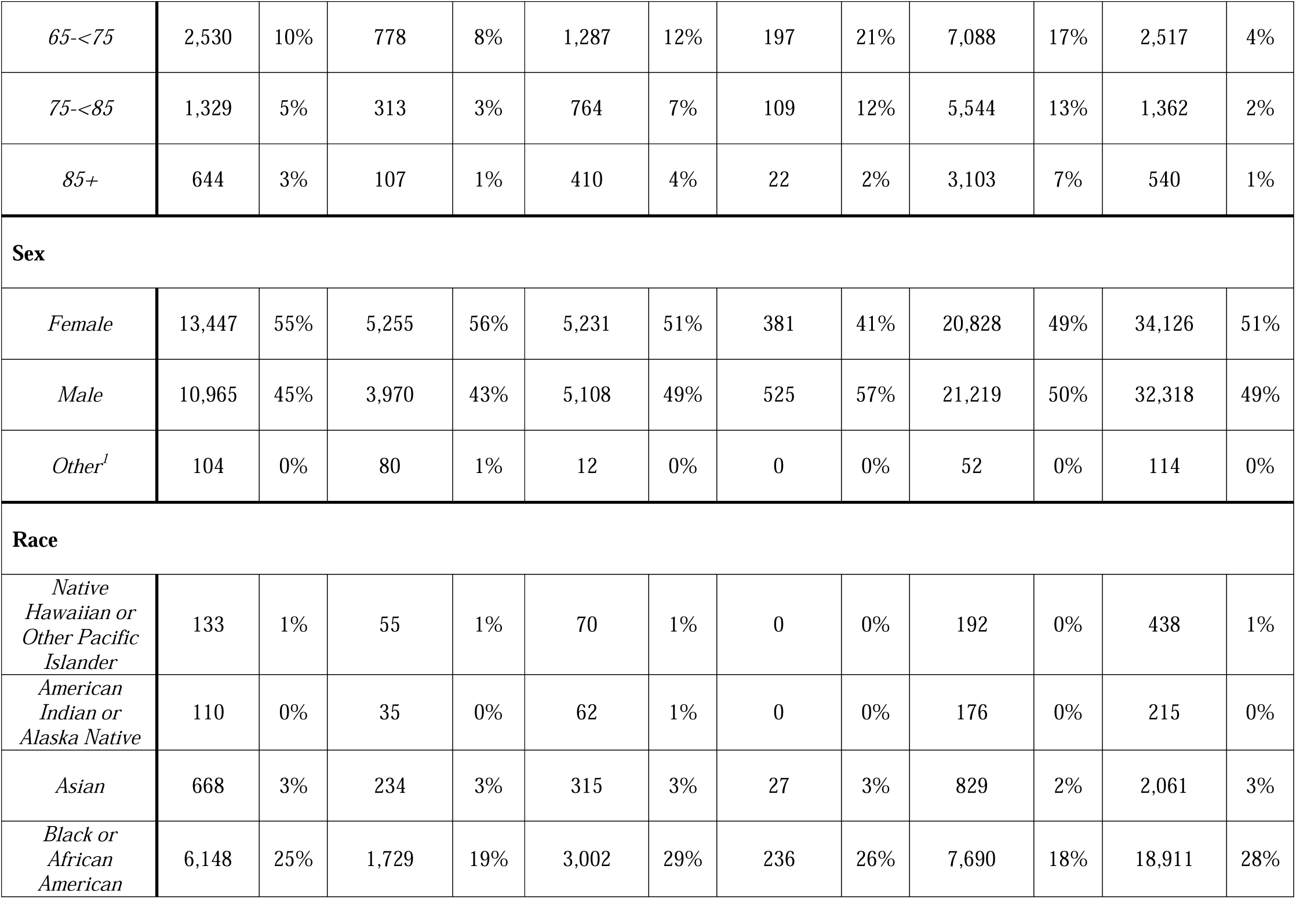
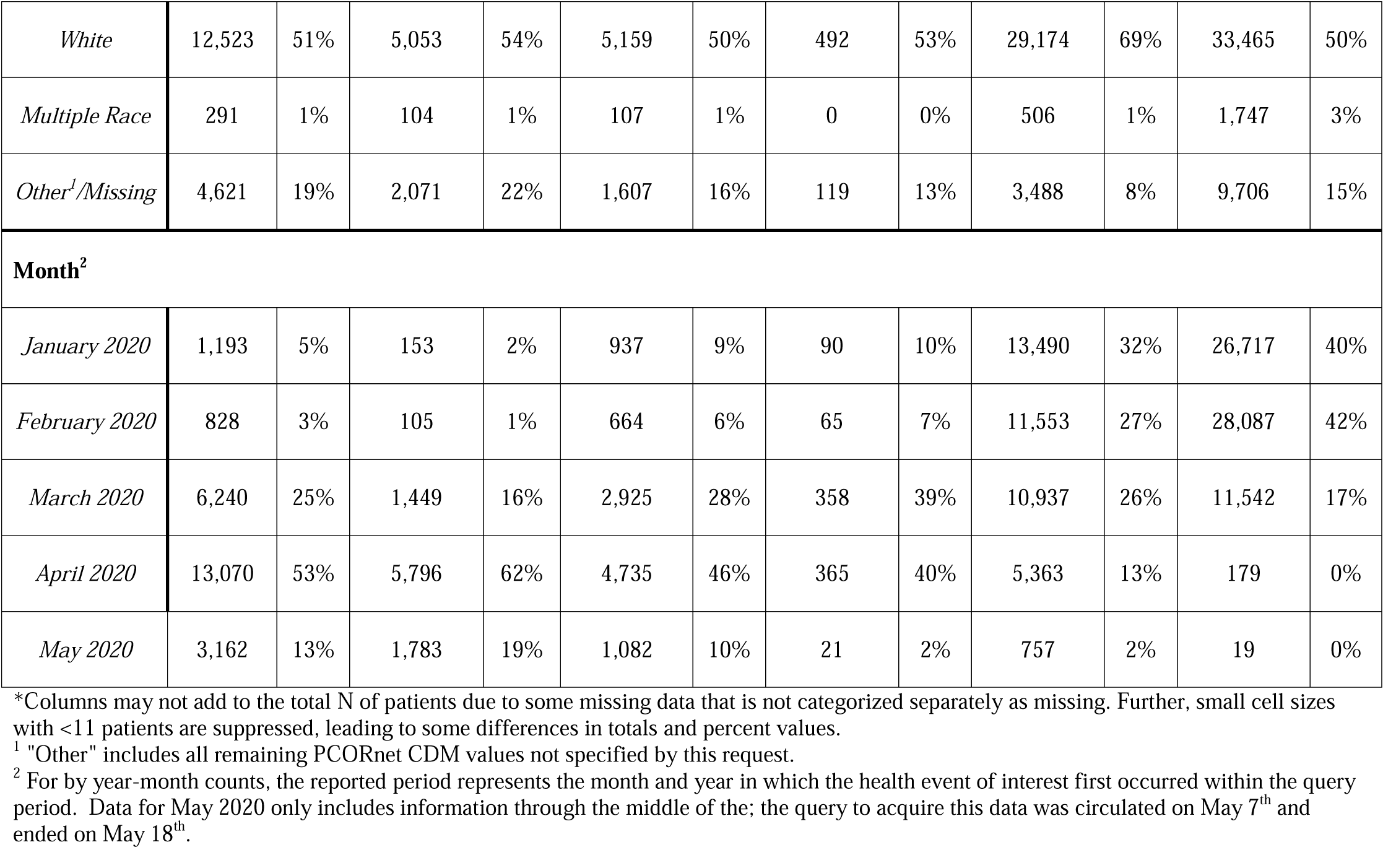
Characteristics of patients diagnosed with COVID-19, viral pneumonia and influenza, January 1 – May 18, 2020, 36 PCORnet Data Contributing Institutions

|  | COVID Diagnosis Any Care Setting |  | COVID Diagnosis in an Ambulatory Setting |  | COVID Diagnosis in an Inpatient or Emergency Department Setting |  | COVID Diagnosis in an Inpatient or Emergency Department Setting AND Mechanical Ventilation |  | Viral Pneumonia Diagnosis AND NO COVID Diagnosis in an Inpatient or Emergency Department Setting |  | Influenza Diagnosis and No COVID Diagnosis in an Inpatient or Emergency Department Setting |  |
| --- | --- | --- | --- | --- | --- | --- | --- | --- | --- | --- | --- | --- |
|  | 24,516 |  | 9,305 |  | 10,351 |  | 921 |  | 42,100 |  | 66,558 |  |
|  | N | % | N | % | N | % | N | % | N | % | N | % |
| **Age** |  |  |  |  |  |  |  |  |  |  |  |  |
| *0-<2* | 1,087 | 4% | 196 | 2% | 770 | 7% | 60 | 7% | 1,968 | 5% | 9,018 | 14% |
| *2-<10* | 976 | 4% | 309 | 3% | 544 | 5% | 44 | 5% | 4,742 | 11% | 24,207 | 36% |
| *10-<20* | 1,105 | 5% | 494 | 5% | 457 | 4% | 33 | 4% | 2,003 | 5% | 8,647 | 13% |
| *20-<45* | 8,890 | 36% | 3,960 | 43% | 2,864 | 28% | 125 | 14% | 6,650 | 16% | 13,173 | 20% |
| *45-<65* | 7,918 | 32% | 3,101 | 33% | 3,249 | 31% | 309 | 34% | 10,966 | 26% | 7,063 | 11% |
| 65-<75 | 2,530 | 10% | 778 | 8% | 1,287 | 12% | 197 | 21% | 7,088 | 17% | 2,517 | 4% |
| 75-<85 | 1,329 | 5% | 313 | 3% | 764 | 7% | 109 | 12% | 5,544 | 13% | 1,362 | 2% |
| 85+ | 644 | 3% | 107 | 1% | 410 | 4% | 22 | 2% | 3,103 | 7% | 540 | 1% |
| **Sex** |  |  |  |  |  |  |  |  |  |  |  |  |
| *Female* | 13,447 | 55% | 5,255 | 56% | 5,231 | 51% | 381 | 41% | 20,828 | 49% | 34,126 | 51% |
| *Male* | 10,965 | 45% | 3,970 | 43% | 5,108 | 49% | 525 | 57% | 21,219 | 50% | 32,318 | 49% |
| *Other<sup>1</sup>* | 104 | 0% | 80 | 1% | 12 | 0% | 0 | 0% | 52 | 0% | 114 | 0% |
| **Race** |  |  |  |  |  |  |  |  |  |  |  |  |
| *Native Hawaiian or Other Pacific Islander* | 133 | 1% | 55 | 1% | 70 | 1% | 0 | 0% | 192 | 0% | 438 | 1% |
| *American Indian or Alaska Native* | 110 | 0% | 35 | 0% | 62 | 1% | 0 | 0% | 176 | 0% | 215 | 0% |
| *Asian* | 668 | 3% | 234 | 3% | 315 | 3% | 27 | 3% | 829 | 2% | 2,061 | 3% |
| *Black or African American* | 6,148 | 25% | 1,729 | 19% | 3,002 | 29% | 236 | 26% | 7,690 | 18% | 18,911 | 28% |
| *White* | 12,523 | 51% | 5,053 | 54% | 5,159 | 50% | 492 | 53% | 29,174 | 69% | 33,465 | 50% |
| *Multiple Race* | 291 | 1% | 104 | 1% | 107 | 1% | 0 | 0% | 506 | 1% | 1,747 | 3% |
| *Other<sup>1</sup>/Missing* | 4,621 | 19% | 2,071 | 22% | 1,607 | 16% | 119 | 13% | 3,488 | 8% | 9,706 | 15% |
| **Month<sup>2</sup>** |  |  |  |  |  |  |  |  |  |  |  |  |
| *January 2020* | 1,193 | 5% | 153 | 2% | 937 | 9% | 90 | 10% | 13,490 | 32% | 26,717 | 40% |
| *February 2020* | 828 | 3% | 105 | 1% | 664 | 6% | 65 | 7% | 11,553 | 27% | 28,087 | 42% |
| *March 2020* | 6,240 | 25% | 1,449 | 16% | 2,925 | 28% | 358 | 39% | 10,937 | 26% | 11,542 | 17% |
| *April 2020* | 13,070 | 53% | 5,796 | 62% | 4,735 | 46% | 365 | 40% | 5,363 | 13% | 179 | 0% |
| *May 2020* | 3,162 | 13% | 1,783 | 19% | 1,082 | 10% | 21 | 2% | 757 | 2% | 19 | 0% |

\*Columns may not add to the total N of patients due to some missing data that is not categorized separately as missing. Further, small cell sizes with <11 patients are suppressed, leading to some differences in totals and percent values.
<sup>1</sup> "Other" includes all remaining PCORnet CDM values not specified by this request.
<sup>2</sup> For by year-month counts, the reported period represents the month and year in which the health event of interest first occurred within the query period. Data for May 2020 only includes information through the middle of the; the query to acquire this data was circulated on May 7<sup>th</sup> and ended on May 18<sup>th</sup>.

Patients with COVID-19 were more likely to have black or African American race (25%) than patients with influenza (18%) or viral pneumonia (15%). Among hospitalized/ED patients, the proportion of patients with black or African American race was similar for COVID-19 and influenza patients (29% and 28%) but higher than for patients with viral pneumonia (18%). However, COVID-19 (16%) and influenza (15%) patients were more commonly categorized as having “missing” or “other” race than those with viral pneumonia (8%).

Comorbidities were common among all infections. Among COVID-19 patients, hypertension was the most common comorbidity (32% for all patients) followed by arrhythmias (20%), pulmonary disease (19%), type 2 diabetes (17%), anemia (14%), and asthma (12%). These diagnoses were more common among COVID-19 patients being treated in the inpatient or emergency department setting (**Fig 2A**). Compared to patients with viral pneumonia treated in the inpatient/ED setting, COVID-19 patients in this same care setting had fewer comorbidities (arrhythmia: 33% vs. 45%; pulmonary disease: 24% vs. 43%; congestive heart failure: 13% vs. 26%; coronary artery disease: 13% vs. 24%); patients with influenza in the inpatient/ED setting had much lower prevalence of comorbidities, likely, in large part, because patients were younger (arrhythmia: 15%; pulmonary disease: 21%; congestive heart failure 4%; coronary artery disease: 4%) (**Fig 2B**). Asthma rates were similar across all three disease groups (14% to 19%). Obesity was more common among COVID-19 patients <65 years old hospitalized or in ED (40% among 20-44-year-olds; 45% among 45-64-year-olds; 36% among 65-74-year-olds) compared to patients with viral pneumonia (32%, 38%, 36%, respectively) and influenza (26%, 37%, 35%, respectively) (**S3 Table**).

**Fig 2.**
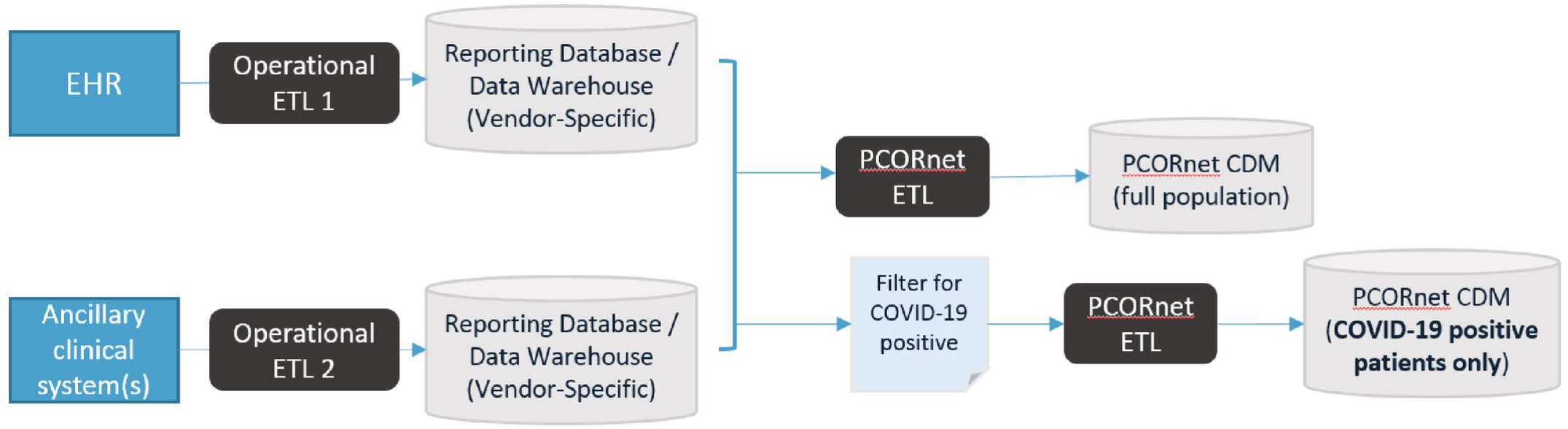
Comorbidities for Patients with COVID-19 Across Care Settings and Compared to Patients with Viral Pneumonia and Influenza in the Inpatient/Emergency Department Setting. These figures show prevalence of various important comorbidities for COVID-19 infections across care settings (A), including all patients and those diagnosed in ambulatory settings, inpatient/emergency department, or using mechanical ventilation. In (B), we compared patients with COVID-19, viral pneumonia, and influenza among those who were hospitalized or in the emergency department. Care settings are not mutually exclusive. Patients could be in all care settings, such as if a patient receives a COVID-19 diagnosis in each of the ambulatory and inpatient/emergency department settings.

Among patients hospitalized or in the ED, critical care procedure/billing codes appeared for 13% of patients diagnosed with COVID-19 compared to 16% of those with viral pneumonia and 2% with influenza. Of treatments assessed, corticosteroids and hydroxychloroquine were used commonly for COVID-19 patients in the inpatient/emergency department setting (12% and 15%, respectively); 10% of patients in this setting used the combination of hydroxychloroquine and azithromycin (**Table 2**). Chloroquine use was very infrequent. Tocilizumab was used most often among patients on mechanical ventilators (11%) as was hydroxychloroquine (33%), with rare use of other anti-interleukin-6 agents. Hydroxychloroquine or the combination of it with azithromycin was used for 1% or fewer of the patients hospitalized or in the ED with viral pneumonia or influenza.

**Table 2:** Treatments used for COVID-19 and Other Infections

|  | **COVID**<br>**Any** |  | **COVID**<br>**Ambulatory** |  | **COVID**<br>**Emergency**<br>**Department**<br>**/Inpatient** |  | **COVID**<br>**Ventilator** |  | **Viral Pneumonia**<br>**Emergency Department**<br>**/Inpatient** |  | **Influenza**<br>**Emergency**<br>**Department**<br>**/Inpatient** |  |
| --- | --- | --- | --- | --- | --- | --- | --- | --- | --- | --- | --- | --- |
| **Hydroxychloroquine** | 1,915 | 8% | 257 | 3% | 1,514 | 15% | 302 | 33% | 618 | 1% | 94 | 0% |
| **Hydroxychloroquine**<br>**+Azithromycin\*** | 1,250 | 5% | 125 | 1% | 993 | 10% | 233 | 25% | 326 | 1% | 26 | 0% |
| **Corticosteroids** | 2,270 | 9% | 354 | 4% | 1,202 | 12% | 198 | 21% | 10,259 | 24% | 5,074 | 8% |
| **Tocilizumab** | 346 | 1% | 12 | 0.001% | 284 | 3% | 101 | 11% | 16 | 0.0004% | 0 | 0% |

\*Not mutually exclusive; patients on hydroxychloroquine and azithromycin are also counted in the hydroxychloroquine row.

## Discussion

In this article, we describe one of the largest cohorts reported to date, using information on 24,516 patients diagnosed with COVID-19 across the U.S. Unlike most other studies, we compare patients across care settings and describe patients diagnosed with viral pneumonia (N=79,639) and influenza (N=163,984) during the COVID-19 pandemic. Comorbidities were common among patients diagnosed with COVID-19, especially those cared for in the emergency department and inpatient setting; prevalence of comorbidities was more higher among patients diagnosed with viral pneumonia and lower among those with influenza. Treatments, such as hydroxychloroquine were commonly used among patients diagnosed with COVID-19. Among patients on mechanical ventilators, 1/3 of patients were using hydroxychloroquine and ¼ using it in combination azithromycin. Obtaining real-world, comprehensive information on a diverse, national population of patients with COVID-19 is critical to help guide policymakers as well as clinical and public health leaders regarding planning for future COVID-19 response. These results provide information on the high burden of chronic disease among COVID-19 patients, compared to those with other similar infections, especially among patients requiring ED visits or hospitalization. The depth and breadth of the PCORnet data infrastructure will facilitate wide-ranging observational and interventional research, with strong generalizability.

Several prior studies have assembled and characterized cohorts of COVID-19 patients mostly using lab test results and extracts from medical records or case reports. These studies originated in China and have emerged from various hotspots across the world as the pandemic has spread. One of the first reports included data on 1099 hospitalized patients across 30 Chinese provinces, representing 14% of patients who had been hospitalized in 552 hospitals at the time. In the cohort, mean age was 47 years, and 24% of patients had an underlying health condition, of which hypertension (15%) and diabetes (7%) were the most common. Patients with more severe disease were more likely to have an underlying health condition (39%) (22). Underscoring the need for large cohorts over diverse geographic regions, subsequent reports from China have found wide variation in characteristics of patients infected with COVID-19 (23). For example, in one study of hospitalized patients across China, patients in Hubei province, the epicenter of the outbreak, were older and had a higher burden of comorbidities (33%) compared to patients outside of Hubei (20%) (24). Diabetes rates were 12% vs. 5%, and hypertension was 24% vs. 12%.

Studies in the United States began with single case reports or case series in Washington state (1, 25) and have grown to include large collections of patients, typically within single healthcare systems or from case reports to public health authorities. In the Northwell healthcare system in New York State, investigators captured data on 5700 inpatients, mostly hospitalized in March 2020, from their EHR data warehouse. Median age of patients was 63 years, 23% were black or African American, and 40% were female; 57% had hypertension, 42% obesity (severe obesity 19%), and 34% diabetes. Among patients, 14% received treatment in an intensive care unit, and 12% had received mechanical ventilation (26).

Research and surveillance networks have begun reporting on COVID-19. The Observational Health Data Sciences and Informatics (OHDSI) group, which manages the Observational Medical Outcomes Partnership (OMOP) Common Data Model, has published data characterizing patients with COVID-19 compared to influenza. OHDSI uses a similar process to PCORnet, with queries that do not require sharing of patient-level data (27). OHDSI network partners contributed information on COVID-19 patients hospitalized in South Korea (captured from a national health insurance claims database) and in three US healthcare systems. Of 6806 patients (75% of them from South Korea) identified based on positive SARS-CoV-2 lab tests, the most common chronic conditions were hypertension (22% to 70% across sites), anemia (14% to 30%), and diabetes (18% to 43%), with the highest burden of chronic disease by far among the 577 patients cared for in Department of Veterans Affairs facilities. In contrast to our study, patients in this OHDSI cohort hospitalized with influenza from 2014 to 2019 were older and had a higher burden of comorbidities than those with COVID-19.

Data on children with COVID-19 are sparse. Among 2572 children under 18 years reported to the Centers for Disease Control and Prevention, 345 children had information on underlying health conditions; 23% of these children had documentation of at least one, most of which were asthma or cardiovascular disease (8). Our study has information available for 3168 children under 20 years of age, 56% of whom were hospitalized; in future iterations of our work, we will have the capacity to report separately on the specific comorbidity burden among children diagnosed with COVID-19 compared to other viral illnesses.

COVID-19 surveillance in PCORnet provides several unique opportunities. The network infrastructure and large samples can allow for creation of research-ready cohorts and in-depth assessment of patients, based on demographic or clinical characteristics. Analyses can be done in a distributed fashion without sharing of patient-level data, including predictive modeling, and aggregation across diverse sites can increase generalizability. Large sample sizes with granular clinical information will be increasingly important to capture information on patient subgroups, defined by demographics or less common presentations or outcomes of COVID-19, such as emerging reports of neurologic complications or rare complications in children (28). PCORnet will also facilitate observational research and clinical trials on various treatments; even in this early phase of data capture, we have identified thousands of patients receiving treatments, such as hydroxychloroquine. By continuously updating and refining COVID-19 data, PCORnet will allow for contemporaneous comparisons to patients diagnosed with other similar illnesses, such as viral pneumonia and influenza in this study as well as concurrent trials; soon, we will compare patients who have “positive” and “negative” SARS-CoV-2 tests. The investment in changing the processes for data management – moving to a weekly update for patients with respiratory illness from a quarterly update – set up the network to make ongoing contributions to the investigation of COVID-19.

In contrast, PCORnet does not allow for representative estimates of overall COVID-19 cases across the U.S. Because PCORnet has a fixed number of institutions and is not designed to be representative of the U.S. population, the prevalence of cases across states in PCORnet is not equivalent to the actual state prevalence of COVID-19 patients. Also, in this early stage of exploring COVID-19 in PCORnet, the case definition depended on diagnostic codes. Patients actively diagnosed with COVID-19, which in our case included codes for other coronaviruses, likely differ from those who test positive for SARS-CoV-2. Diagnostic codes are less sensitive than lab test results though perhaps more specific, considering that many patients with positive lab tests are asymptomatic and do not have the syndrome of COVID-19. Sensitivity of diagnostic codes for COVID-19 was likely much lower for patients who were diagnosed before an ICD-10-CM code was available for COVID-19 (April 1, 2020) (29). Prior to that time, the Centers for Disease Control and Prevention provide interim guidance on use of existing coronavirus codes that were not specific to COVID-19 (30). The presence of the U07.1 code already has led to more case identification, with most of the cases identified in April and May using this code. Because of the uncertainty of diagnostic codes in the early phase of the pandemic, some patients with other coronavirus infections would have been misclassified as having COVID-19 in our study.

### Limitations

Several limitations are inherent to working with structured healthcare data. First, electronic health records may not have complete data on some patients, especially patients diagnosed in virtual visits or who were diagnosed and treated entirely outside of a PCORnet health system, including in skilled nursing facilities or nursing homes. In such cases, the PCORnet Common Data Model may not have any information about a COVID-19 infection or inadequate information on chronic diseases and medication use. We separately captured information on patients who were seen in the healthcare system prior to their hospitalization or emergency department visit for COVID-19; among these patients, prevalence of comorbidities was slightly higher when compared to all patients seen in the emergency department or hospitalized. Missing data can also be an issue for variables such as race. Some medications being used in research protocols also were likely missing because they are not captured in routine medication administration tables. Second, because the research network was set up to accommodate quarterly refreshes of clinical data, we quickly changed protocols to allow for weekly updates. The standard quarterly data refreshes undergo detailed curation to ensure that data are compliant with standards of the Common Data Model, and such efforts are not possible with weekly refreshes. Erroneous records may exist that will only be identified later through data curation. The PCORnet Coordinating Center will soon implement regular data curation for the COVID-19 Common Data Model. Third, during these initial queries, we categorized weight status by age group, but we did not fully separate children or adults for the determination of comorbidities and treatments; this led to lower prevalence of comorbidities for patients with influenza. Future queries will separate categorize adults and children. Fourth, overlap likely existed between disease groups. Patients diagnosed with viral pneumonia or influenza might have had COVID-19 but either did not receive a test or were misclassified based on diagnostic coding. Only 12% of patients with viral pneumonia and 1% with influenza had a recorded SARS-CoV-2 test, compared to 68% for patients with COVID-19. These SARS-CoV-2 test results are likely an undercount; some sites have not yet fully loaded their test results into PCORnet data models, and others might not have information on tests done outside of the health system, such as in drive-thru testing facilities or private labs.

### Conclusion and Next Steps in Development

PCORnet has captured information on one of the largest cohorts to date of patients diagnosed with COVID-19 and other respiratory illnesses. Patients diagnosed with COVID-19 have a high burden of chronic disease, and treatments are common. We have updated data infrastructure to allow for weekly updates and queries of patients, with COVID-19 case definitions initially based on diagnostic codes. We will soon identify and characterize cohorts based on lab results, along with additional improvements such as identifying patients based on institutional registries, improved definitions of care settings, and detailed stratifications by patient characteristics.

### Collaborative Authors: PCORnet^®^ Network Partners

H. Timothy Bunnell, PhD, Nemours Children’s Health System; Ryan Carnahan, PharmD, MS, The University of Iowa College of Public Health, Department of Epidemiology; Alanna M. Chamberlain, PhD, MPH, Department of Health Sciences Research, Mayo Clinic; Dimitri A Christakis, MD MPH, Center for Child Health, Behavior & Development, Seattle Childrens Research Institute, University of Washington; Lindsay G. Cowell, MS, PHD, UT Southwestern Medical Center; Janis L. Curtis, MSPH, MA, Duke University School of Medicine; Julia A. Fearrington, Harvard Pilgrim Health Care Institute; Soledad A. Fernandez, PhD, The Ohio State University; David A Hanauer, MD, MS, University of Michigan; Kristin Herman, Tallahassee Memorial Healthcare, Inc; Rachel Hess, MD, MS, Departments of Population Health Sciences and Internal Medicine, University of Utah School of Medicine; Philip Giordano, MD, Orlando Health, Inc.; Wenke Hwang, PhD, Penn State University College of Medicine; Adam Lee, MBA, NC TraCS Institute, University of North Carolina, Chapel Hill; Harold Lehmann MD PhD, Health Science Informatics, Johns Hopkins School of Medicine; Simon Lin, MD, MBA, Nationwide Children’s Hospital; Scott Mackey, DO, LCMC Health; Narayana Mazumber, MS, Allina Health; James McClay, MD, MS, FACEP, FAMIA, University of Nebraska Medical Center; Abu Saleh Mohammad Mosa, PhD, MS, FAMIA, University of Missouri School of Medicine; Jihad S. Obeid, MD, FAMIA, Biomedical Informatics Center, Department of Public Health Sciences, Medical University of South Carolina; Brian Ostasiewski, B.S., Wake Forest School of Medicine; Nathan M. Pajor, MD, MS, Division of Pulmonary Medicine, Cincinnati Children’s Hospital Medical Center, and Departments of Pediatrics, University of Cincinnati; Aruradha Paranjape, MD, MPH, Lewis Katz School of Medicine at Temple University; Suchitra Rao, MBBS, MSCS, University of Colorado School of Medicine and Children’s Hospital Colorado; Pedro Rivera, OCHIN; Patricia S. Robinson, PhD, APRN, NE-BC, AdventHealth; Marc Roseman, Ann & Robert H. Lurie Children’s Hospital of Chicago; Steffani Roush, Marshfield Clinic Health System; Jonathan C. Silverstein, MD, MS, Department of Biomedical Informatics, University of Pittsburgh School of Medicine; Alexander Stoddard, MS,CTSI, Medical College of Wisconsin; William E. Trick, Center for Health Equity & Innovation, Cook County Health; Katherine J Worley, MLIS, Vanderbilt University Medical Center

## Data Availability

All code lists and query tools are available publicly on GitHub (cited in manuscript).
Source data for this study are kept behind institutional firewalls and are subject to
sharing in accord with the PCORnet Data Security Agreement and Data Use Agreements. The PCORnet Steering Committee and individual data contributing sites will need to approve of release of any data.

https://github.com/PCORnet-DRN-OC/Query-Details/tree/master/COVID_weekly_query/2020_04_29

## Abbreviations

EHR: electronic health record
CDM: Common Data Model
ETL: extract-transfer-load procedure to move data from a clinical data warehouse to the PCORnet Common Data Model
HTN: hypertension
Diabetes: Type 2 Diabetes Mellitus
Pulm: pulmonary disease
CKD: chronic kidney disease
CAD: coronary artery disease
CHF: congestive heart failure
Coag: Coagulopathy
Cancer: Cancer other than non-melanoma skin cancer
Immunosupp: use of immunosuppressive medications
Any: any care setting
Amb: ambulatory care setting
IP/ED: inpatient or emergency department
Vent: inpatient or emergency department with procedure codes indicating use of mechanical ventilation
PNA: pneumonia

## Acknowledgement

The Clinical Research Networks, Health Plan Research Networks, and Coordinating Center reported in this publication are Network Partners in PCORnet, the National Patient-Centered Clinical Research Network. PCORnet has been developed with funding from the Patient-Centered Outcomes Research Institute (PCORI). The Network Partners’ participation in PCORnet is funded through the following PCORI Awards: Coordinating Center (PCORI-CC2-Duke-2016); ADVANCE (RI-CRN-2020-001); CaPriCORN (RI-CRN-2020-002); GPC (RI-CRN-2020-003); HealthCore (HP-1510-32545); Insight (RI-CRN-2020-004); OneFlorida (RI-CRN-2020-005); PaTH (RI-CRN-2020-006); PEDSnet (RI-CRN-2020-007); PRACnet (HP-1511-32713); REACHnet (RI-CRN-2020-008); and, STAR (RI-CRN-2020-009). Authors report no competing interests associated with this publication.

